# Cost-Effectiveness of Ensitrelvir for COVID-19 Post-Exposure Prophylaxis Among High-Risk Household Contacts in Japan: A Model-Based Cost-Utility Analysis

**DOI:** 10.64898/2026.09.15.26361876

**Authors:** Toshibumi Taniguchi, Misuzu Yahaba, Hiroshi Yoshikawa, Hidetoshi Igari, Daisuke Sato

**Author notes:** Corresponding author Toshibumi Taniguchi, MD, PhD, Department of Infectious Diseases, Chiba University Hospital, 1-8-1 Inohana, Chuo-ku, Chiba 260-8677, Japan.

## Abstract

**Background:** Ensitrelvir reduced symptomatic COVID-19 after household exposure in the SCORPIO-PEP trial, but its economic value as post-exposure prophylaxis is unknown.

**Objective:** To estimate the cost-effectiveness of ensitrelvir post-exposure prophylaxis for high-risk household contacts in Japan.

**Methods:** We developed a decision-analytic cost-utility model from the Japanese public healthcare payer perspective. Trial risks of symptomatic COVID-19 through day 10 were linked to Japanese evidence on hospitalization, mortality, costs, quality of life, and life expectancy. Because the trial observed no COVID-19-related hospitalization or death, downstream outcomes were modeled only through prevention of symptomatic disease. We performed deterministic, probabilistic, structural, subgroup, and threshold analyses.

**Results:** Ensitrelvir increased costs by JPY 54,945 and gained 0.003267 QALYs per exposed contact, yielding an ICER of JPY 16.82 million/QALY. None of 10,000 simulations was cost-effective at JPY 5 million/QALY. Applying the overall-trial relative risk to the high-risk baseline yielded JPY 19.24 million/QALY. Restricting post-acute utility loss to 6 months, halving it, or excluding it yielded ICERs of JPY 23.62 million, JPY 26.76 million, and JPY 65.51 million/QALY, respectively. The drug acquisition-cost threshold was JPY 11,021 per course.

**Conclusions:** At current Japanese costs, ensitrelvir post-exposure prophylaxis for high-risk household contacts was unlikely to be cost-effective at JPY 5 million/QALY. This conclusion remained unfavorable under conservative efficacy, post-acute utility, hospitalization-attribution, and mortality assumptions. Severe-outcome benefits remain model-based extrapolations rather than trial-observed effects.

**Key Points for Decision Makers:**

- The pivotal post-exposure prophylaxis trial showed a substantial reduction in symptomatic COVID-19, but no COVID-19-related hospitalization or death occurred; modeled severe-outcome benefits therefore depend on external Japanese evidence.
- At current Japanese drug and delivery costs, the base-case ICER was JPY 16.82 million per QALY. The probability of cost-effectiveness was 0% at JPY 5 million per QALY and 2.28% at JPY 10 million per QALY.
- The acquisition-cost threshold was approximately JPY 11,000 per course. Conservative efficacy and post-acute utility scenarios remained above JPY 19 million and JPY 23 million per QALY, respectively.

## 1 Introduction

Household exposure remains an important setting for SARS-CoV-2 transmission because contact intensity is high and prophylaxis must be initiated quickly. Ensitrelvir is an oral 3C-like protease inhibitor administered for 5 days. In SCORPIO-PEP, treatment initiated within 72 hours of symptom onset in the index case reduced symptomatic COVID-19 through day 10. A prespecified subgroup analysis among participants with at least one risk factor for severe COVID-19 found symptomatic disease in 9 of 382 ensitrelvir recipients and 37 of 374 placebo recipients [1].

Japan approved ensitrelvir for prevention after household exposure in March 2026. The approved course comprises three 125-mg tablets on day 1 and one tablet daily on days 2-5, for a total of seven tablets [2, 3]. The availability of an oral post-exposure prophylaxis creates a policy question that differs from treatment after diagnosis: the intervention cost is incurred for every exposed contact, whereas serious outcomes are uncommon and were not observed in the pivotal trial.

Japanese economic evaluations typically report costs and quality-adjusted life-years (QALYs) from a public healthcare payer perspective and examine uncertainty around model structure and extrapolation [4]. Economic evaluations should also distinguish trial-observed outcomes from outcomes projected using external data, particularly when the event pathway includes rare hospitalization and death. We followed the Consolidated Health Economic Evaluation Reporting Standards 2022 (CHEERS 2022) [5].

No published cost-utility analysis of ensitrelvir post-exposure prophylaxis was identified in the targeted literature searches documented in Online Resource 1. We therefore developed a transparent, reproducible decision-analytic model to estimate its cost-effectiveness among high-risk household contacts in Japan, quantify the contribution of acute and post-acute outcomes, and identify price and risk thresholds relevant to targeted implementation.

## 2 Methods

### 2.1 Study Design and Decision Problem

We compared ensitrelvir post-exposure prophylaxis with no pharmacologic prophylaxis among household contacts with at least one risk factor for severe COVID-19. The analysis adopted the Japanese public healthcare payer perspective. Drug acquisition and prophylaxis-delivery inputs were valued using 2026 prices and fee schedules. Transferred outpatient and inpatient cost estimates retained their 2023 and 2021 source-year nominal values because no defensible item-level re-pricing method was available; 0.8-1.2 cost multipliers were used to test price-year and practice-pattern uncertainty. Outcomes were expressed in QALYs, and incremental net monetary benefit (INMB) used a reference value of JPY 5 million per QALY. Future QALYs were discounted at 2% annually. The acute model began at the prophylaxis decision and projected downstream lifetime QALY loss from acute death. Post-acute morbidity was modeled for 1 year after symptomatic infection.

### 2.2 Population, Intervention, and Comparator

The target population reflected the prespecified high-risk subgroup analysis of SCORPIO-PEP: SARS-CoV-2-negative household contacts of an index case who were eligible to start prophylaxis within 72 hours after index-case symptom onset [1]. Ensitrelvir was modeled as a 5-day course using the Japanese approved regimen [2, 3]. The comparator was no pharmacologic prophylaxis. Standard clinical care after any breakthrough infection was assumed to be available in both strategies.

### 2.3 Model Structure and Causal Assumptions

The decision tree is shown in Fig. 1. Each strategy led to symptomatic or no symptomatic COVID-19. Symptomatic cases could remain outside hospital or experience hospitalization attributable to COVID-19, followed by survival or in-hospital death. Surviving symptomatic cases incurred acute utility loss and a 1-year post-acute utility profile. Ensitrelvir was not assigned a direct effect on hospitalization, mortality, or post-acute burden among breakthrough cases.

**Fig. 1.**
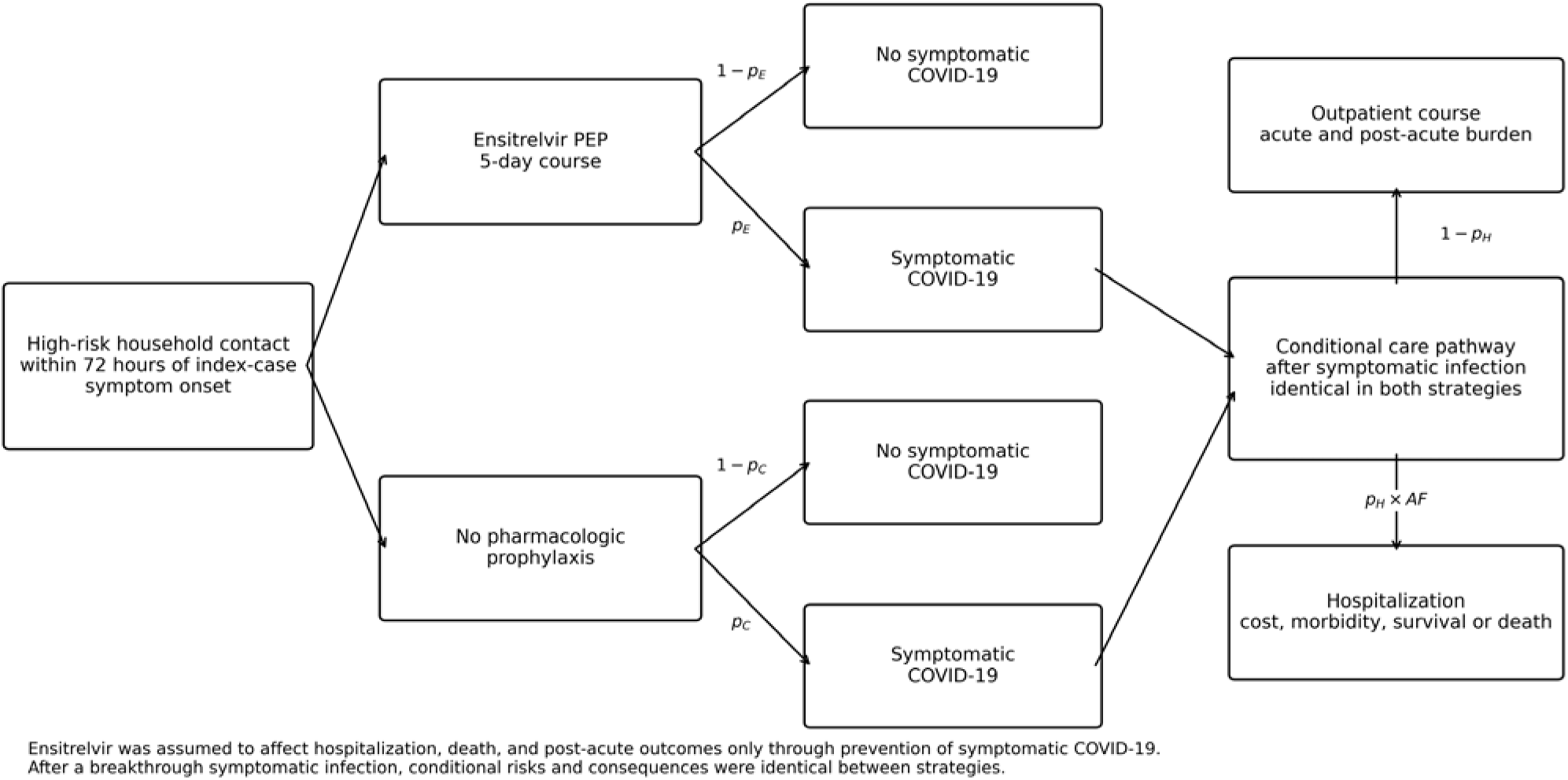
Decision-analytic model structure. Abbreviations: AF attribution fraction; PEP post-exposure prophylaxis. pE and pC are the probabilities of symptomatic COVID-19 with ensitrelvir PEP and no pharmacologic prophylaxis, respectively; pH is the observed hospitalization probability conditional on symptomatic COVID-19

This mediated-effect assumption was selected because SCORPIO-PEP observed no COVID-19-related hospitalization or death in either arm [1]. Conditional risks and consequences after symptomatic COVID-19 were therefore held equal between strategies. The base-case hospitalization source measured 28-day all-cause admission. A separate attribution fraction mapped this endpoint to hospitalizations assigned COVID-19 cost and mortality. The base case used 100% attribution, with 50% and 0% structural analyses.

### 2.4 Clinical Effectiveness

Arm-specific symptomatic risks were derived directly from the high-risk trial subgroup: 2.356% with ensitrelvir and 9.893% with placebo [1]. The absolute risk reduction was 7.537 percentage points and the number needed to treat to prevent one symptomatic case was 13.27. Because subgroup efficacy may be less stable than the overall trial estimate, a conservative scenario applied the overall-trial relative risk of 0.33 to the high-risk untreated risk. Trial adverse-event rates were similar between groups; incremental adverse-event cost and QALY loss were therefore zero in the base case and varied in deterministic sensitivity analysis.

### 2.5 Hospitalization and Mortality

The conditional hospitalization probability was 0.785%, based on 28-day all-cause hospitalization among untreated Japanese high-risk outpatients with diagnosed COVID-19 [6]. The observed endpoint was not restricted to admissions causally attributable to COVID-19, which motivated the explicit attribution parameter. In-hospital mortality was 12%, based on a separate contemporary Japanese inpatient database that included the late Omicron period [7]. These sources differed in age, insurance coverage, treatment status, and care setting. Death outside hospital was set to zero in the publication base case. To examine post-diagnosis treatment consistency, an exploratory scenario assumed that 50% of breakthrough cases received ensitrelvir treatment, added treatment cost to symptomatic care, and reduced hospitalization risk using the treated and untreated risks reported in the same claims study.

Remaining QALYs lost per acute death were calculated from the 2025 Japanese abridged life table and age- and sex-specific Japanese EQ-5D-5L population norms [10, 11]. The proxy cohort used age 82 years and 54% male, based on the contemporary hospitalized cohort, and produced 6.469 discounted QALYs lost per death. Age-75 and age-85 calibrations were evaluated as scenarios.

### 2.6 Health-Related Quality of Life

The acute outpatient utility decrement was 0.258, calculated as the difference between symptomatic test-negative controls and COVID-19 cases in a Japanese Omicron-period EQ-5D-5L study [8]. The decrement was applied for 7 days. Hospitalized cases received a broader proxy decrement during a 21-day stay. No additional acute decrement was assigned from day 8 through day 30, a conservative assumption that favors no prophylaxis.

Post-acute QALY loss was integrated from a retrospective online recall survey of adults tested at a single Tokyo clinic, with 302 COVID-19 respondents and 77 test-negative controls [8]. Participants retrospectively reported EQ-5D-5L for the acute period and months 1-3, 3-6, and 6-12. The base-case area between the group profiles was 0.03225 QALYs per surviving symptomatic case. Because the survey had limited controls, no prospectively measured baseline utility, and potential recall and response-selection bias, conservative scenarios excluded the 6-12-month interval, halved the integrated loss, or removed post-acute utility entirely. We did not replace the symptomatic test-negative comparator with general-population norms because that would not preserve like-for-like symptom and selection characteristics. A separate Japanese population study was used only for contextual and legacy scenario checks [15].

### 2.7 Costs

The acquisition cost was JPY 49,630 per course, calculated from the listed price of JPY 7,090 per tablet and seven tablets [2]. The base delivery cost was JPY 8,100, microcosted from the FY2026 Japanese medical and dispensing fee schedules [12]. It included a face-to-face initial consultation, qualitative antigen testing, specimen collection, test interpretation, prescription, and representative pharmacy fees. The FY2026 two-point outpatient inflation add-on, equivalent to JPY 20, was not included because its effect is immaterial. Facility-dependent wage add-ons were excluded because eligibility varies. Face-to-face and telemedicine pathways without separately billed testing were evaluated in scenarios.

Outpatient symptomatic care cost JPY 26,927 per case, based on a prior Japanese COVID-19 economic model using 2023 values [13]. Hospitalization cost JPY 1,304,431 per admission, based on a 2021 Japanese hospital cost study [9]. These transferred source-year values were not relabeled as 2026 prices; deterministic cost multipliers tested the potential effect of fee revisions, inflation, and practice-pattern differences. Because a transportable Japanese estimate of incremental post-acute direct medical cost was not identified, this cost was set to zero in the payer base case and varied from JPY 50,000 to JPY 200,000 per surviving symptomatic case.

### 2.8 Analyses

We calculated expected cost, QALY loss, symptomatic cases, hospitalization, death, incremental cost, incremental QALYs, ICER, INMB, and events prevented per exposed contact and per 1,000 contacts. Cost-effectiveness was evaluated against JPY 5 million per QALY. Costs and QALYs were reported without rounding in the computational outputs and rounded for presentation.

One-way sensitivity analysis used prespecified parameter ranges and ranked parameters by their effect on INMB. The main tornado figure excluded drug acquisition cost because price was evaluated separately by value-based threshold analysis; the full deterministic table retained all parameters. In probabilistic sensitivity analysis, the untreated symptomatic risk was sampled from a Jeffreys beta posterior and the published relative risk from a log-normal distribution derived from its 95% confidence interval. Because their covariance was unavailable, independence was assumed. Other probabilities and utilities used beta or PERT-type distributions, costs used gamma or PERT-type distributions, and fixed structural parameters were excluded from probabilistic sampling. PERT-type distributions were used only when lower, modal, and upper bounds were available without an empirical parametric fit. We ran 10,000 simulations with seed 20260825.

Structural analyses excluded acute mortality benefit, excluded or reduced post-acute health loss, varied the hospitalization attribution fraction, modified delivery pathways, incorporated post-diagnosis antiviral treatment, and varied post-acute costs. An efficacy scenario applied the overall-trial relative risk to the high-risk untreated risk. Exploratory risk scenarios used observed older-patient risk and hospitalization multipliers for immunocompromised conditions reported in Japanese data [14]. Two static bounding analyses added 0.2 or 0.5 downstream symptomatic cases prevented per directly prevented case. These calculations assigned downstream cases the same conditional hospitalization and post-acute outcomes as the high-risk contact population and therefore represent upper bounds. They are not dynamic transmission models and were excluded from the base case and probabilistic analysis. Threshold analyses solved for zero INMB across drug cost, total delivery cost, hospitalization risk, post-acute QALY loss, and other uncertain inputs.

### 2.9 Verification, Input Traceability, and Reproducibility

Model verification followed good-practice principles for decision-analytic models [16, 17]. Automated checks verified parameter ranges, source links, trial arithmetic, scenario execution, and reproducibility. Independent audit scripts recalculated the PEP microcost, post-acute QALY integral, and mortality-QALY estimate from source tables. Unit tests assessed event arithmetic, attribution logic, intervention costs, life-table calculations, downstream-case logic, and probabilistic reproducibility. Comparisons with registered source values were treated as input traceability and face-validity checks, not as independent predictive external validation.

All numeric inputs, evidence sources, extrapolations, scenario overrides, code, outputs, and environment information are supplied in the Online Resources. The exact public release of model version 0.3.2 is available at https://github.com/toshtanig/ensitrelvir_PEP_CEA, tagged as v0.3.2, and will be archived in a permanent repository upon journal submission. Online Resource 2 is a machine-readable workbook, and Online Resource 3 contains the exact archived release used for this analysis.

### 2.10 Patient and Stakeholder Involvement

Patients and members of the public were not directly involved in model development. The decision problem was framed around payer and policy decisions. The absence of direct stakeholder involvement is reported as a limitation and should be addressed in subsequent implementation research.

### 2.11 Use of Generative Artificial Intelligence

OpenAI GPT-5.6 Pro was used to assist with drafting, document formatting, and code review. Numerical outputs were generated by the versioned Python model. The authors are responsible for independently verifying all analyses, source citations, interpretation, and final wording before submission.

**Table 1.** Key model inputs.

| Parameter | Base case | Deterministic range or trial basis | Source |
| --- | --- | --- | --- |
| Symptomatic COVID-19, ensitrelvir | 0.02356 | 9/382 observed | Trial [1] |
| Symptomatic COVID-19, no prophylaxis | 0.09893 | 37/374 observed | Trial [1] |
| Hospitalization conditional on symptomatic COVID-19 | 0.00785 | 0.00494-0.01298 | Japanese claims [6] |
| Hospitalization attribution fraction | 1.00 | 0.00-1.00 | Structural assumption |
| In-hospital mortality | 0.12 | 0.06-0.20 | Japanese inpatient cohort [7] |
| Outpatient acute utility decrement | 0.258 | 0.181-0.335 | Japanese retrospective recall EQ-5D-5L survey [8] |
| Post-acute QALY loss per surviving symptomatic case | 0.03225 | 0.01067-0.052 | Japanese retrospective recall EQ-5D-5L survey [8] |
| Remaining QALYs lost per acute death | 6.469 | 5.321-9.779 | Life-table calibration [7, 10, 11] |
| Ensitrelvir acquisition cost, JPY | 49,630 | 0-49,630 | Official price [2] |
| PEP delivery cost, JPY | 8,100 | 4,530-10,000 | FY2026 microcost [12] |
| Outpatient symptomatic-care cost, JPY | 26,927 | 21,542-32,312 | Japanese economic model [13] |
| Hospitalization cost, JPY | 1,304,431 | 798,810-6,210,607 | Japanese hospital study [9] |
| Post-acute direct medical cost, JPY | 0 | 0-200,000 | Evidence-gap assumption |

Abbreviations: COVID-19 coronavirus disease 2019; EQ-5D-5L EuroQol 5-Dimension 5-Level; FY fiscal year; JPY Japanese yen; PEP post-exposure prophylaxis; QALY quality-adjusted life-year.
The third column reports either the deterministic range or the trial n/N basis for directly observed risks; these entries are not uniformly confidence intervals.

## 3 Results

### 3.1 Base-Case Results

Per exposed contact, ensitrelvir cost JPY 58,601 and generated 0.001021 QALY loss, whereas no prophylaxis cost JPY 3,656 and generated 0.004288 QALY loss. The incremental cost was JPY 54,945 and the QALY gain was 0.003267, producing an ICER of JPY 16.82 million per QALY. INMB at JPY 5 million per QALY was JPY -38,609.

Among 1,000 exposed contacts, the model projected 75.37 symptomatic COVID-19 cases, 0.592 COVID-19-attributable hospitalizations, and 0.0710 acute deaths prevented. The number needed to treat to prevent one symptomatic case was 13.27. These hospitalization and death effects were extrapolated through the modeled pathway and were not observed in the trial.

### 3.2 Cost and QALY Decomposition

The prophylaxis intervention added JPY 57,730 per contact and reduced acute healthcare cost by JPY 2,785, yielding the net incremental cost of JPY 54,945. Of the total QALY gain, 0.000379 (11.6%) arose from avoided acute morbidity, 0.000459 (14.1%) from modeled mortality avoidance, and 0.002428 (74.3%) from avoided post-acute morbidity.

**Table 2.** Base-case cost-effectiveness results.

| Outcome | Base-case result | Interpretation |
| --- | --- | --- |
| Incremental cost, JPY/contact | 54,945 | PEP cost minus avoided healthcare cost |
| Incremental QALYs/contact | 0.003267 | Positive value favors ensitrelvir |
| ICER, JPY/QALY | 16,817,443 | Incremental cost divided by QALY gain |
| INMB at JPY 5 million/QALY, JPY/contact | -38,609 | Negative value not cost-effective |
| Symptomatic cases prevented/1,000 | 75.37 | Trial-mediated outcome |
| Hospitalizations prevented/1,000 | 0.592 | Model extrapolation |
| Deaths prevented/1,000 | 0.0710 | Model extrapolation |
| Number needed to treat | 13.27 | To prevent one symptomatic case |

Abbreviations: ICER incremental cost-effectiveness ratio; INMB incremental net monetary benefit; JPY Japanese yen; PEP post-exposure prophylaxis; QALY quality-adjusted life-year.
*ICER incremental cost-effectiveness ratio, INMB incremental net monetary benefit, PEP post-exposure prophylaxis.*

### 3.3 Deterministic Sensitivity Analysis

Among non-price inputs, post-acute QALY loss had the largest effect on INMB (Fig. 2), followed by post-acute direct medical cost, untreated symptomatic risk, the relative risk of symptomatic disease, and additional access cost. Drug acquisition cost was intentionally omitted from the main tornado figure because its full 0-100% variation would dominate the display and is more directly interpreted through the separate value-based price threshold. The complete deterministic sensitivity table, including acquisition cost, is supplied in Online Resource 2. No clinical input within its prespecified range reversed the base-case conclusion at the current acquisition price.

**Fig. 2.**
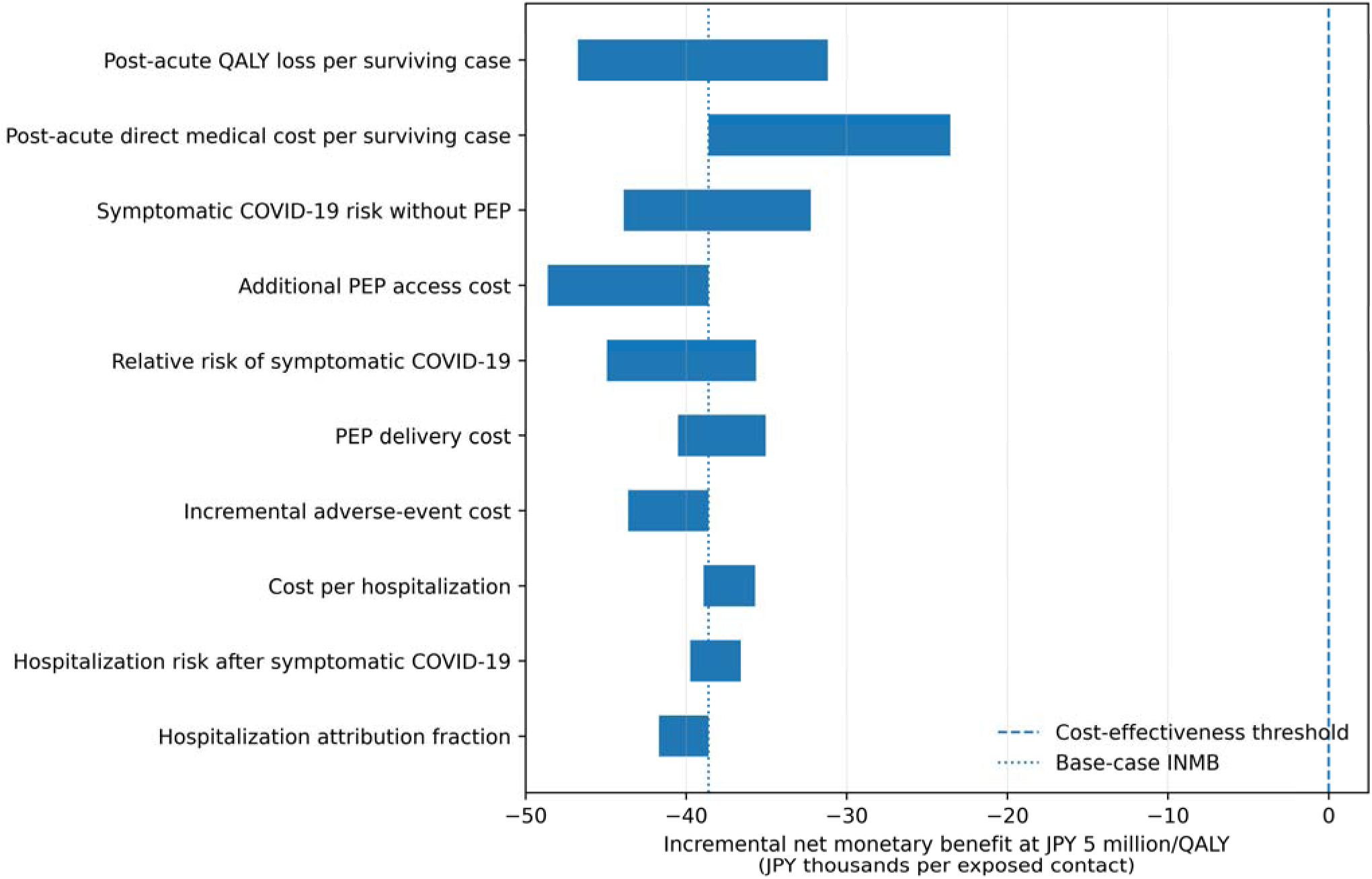
One-way sensitivity analysis ranked by the effect of non-price inputs on incremental net monetary benefit at JPY 5 million per QALY. Drug acquisition cost was excluded from this display and evaluated by a separate value-based price threshold. Abbreviations: INMB incremental net monetary benefit; PEP post-exposure prophylaxis; QALY quality-adjusted life-year

### 3.4 Probabilistic Sensitivity Analysis

Across 10,000 simulations, mean incremental cost was JPY 54,197 (95% simulation interval JPY 51,161-56,659) and mean incremental QALYs were 0.003280 (0.001731-0.005231). All simulations produced a QALY gain, none was cost-saving, and none was cost-effective at JPY 5 million per QALY. The probability of cost-effectiveness was 0.02% at JPY 7.5 million per QALY and 2.28% at JPY 10 million per QALY. The cost-effectiveness acceptability curve is shown in Fig. 3.

**Fig. 3.**
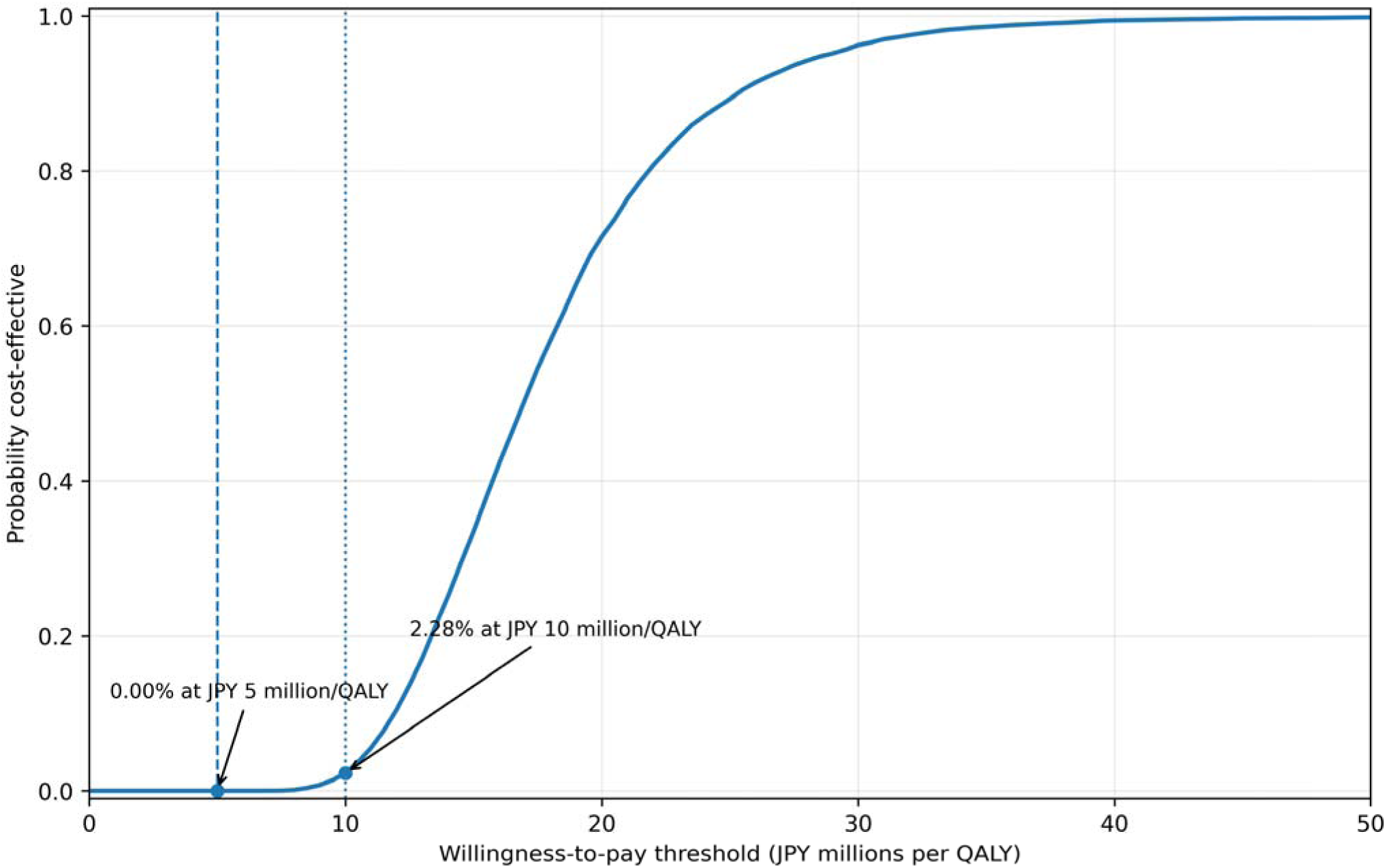
Cost-effectiveness acceptability curve from 10,000 probabilistic simulations. The probabilities of cost-effectiveness were 0% at JPY 5 million per QALY and 2.28% at JPY 10 million per QALY. Abbreviation: QALY quality-adjusted life-year

### 3.5 Structural, Risk, and Threshold Analyses

**Table 3.** Selected scenario analyses.

| Scenario | Incremental cost, JPY | Incremental QALYs | ICER, JPY million/QALY |
| --- | --- | --- | --- |
| Base case | 54,945 | 0.003267 | 16.82 |
| Overall-trial RR 0.33 applied to high-risk baseline | 55,280 | 0.002873 | 19.24 |
| No acute mortality benefit | 54,945 | 0.002810 | 19.55 |
| Post-acute utility through 6 months only | 54,945 | 0.002326 | 23.62 |
| Post-acute utility loss halved | 54,945 | 0.002053 | 26.76 |
| No post-acute utility benefit | 54,945 | 0.000839 | 65.51 |
| 50% breakthrough antiviral treatment uptake | 53,226 | 0.003181 | 16.73 |
| No hospitalization attributed to COVID-19 | 55,701 | 0.002803 | 19.87 |
| 0.2 downstream symptomatic cases per direct case | 54,388 | 0.003921 | 13.87 |
| 0.5 downstream symptomatic cases per direct case | 53,552 | 0.004901 | 10.93 |
| Conditional hospitalization risk 10% | 46,072 | 0.007672 | 6.00 |

Abbreviations: ICER incremental cost-effectiveness ratio; JPY Japanese yen; QALY quality-adjusted life-year; RR relative risk. The 10% conditional hospitalization risk is a hypothetical value not observed in the Japanese sources used in the model. Downstream-case scenarios assign high-risk conditional outcomes and therefore represent upper-bound static calculations.

All prespecified and additional conservative scenarios remained above JPY 5 million/QALY. Risk multipliers are exploratory external extrapolations. The 10% conditional hospitalization scenario is hypothetical and was not observed in any Japanese source used in the model. The downstream-case multipliers are static upper-bound calculations that assign the same high-risk conditional outcomes to downstream cases.

Applying the overall-trial relative risk of 0.33 to the high-risk untreated risk increased the ICER to JPY 19.24 million per QALY. Excluding the 6-12-month post-acute interval, halving post-acute QALY loss, or removing it entirely produced ICERs of JPY 23.62 million, JPY 26.76 million, and JPY 65.51 million per QALY, respectively. Excluding modeled acute mortality produced JPY 19.55 million per QALY, and attributing none of the claims-based admissions to COVID-19 produced JPY 19.87 million per QALY. Incorporating 50% breakthrough-treatment uptake produced JPY 16.73 million per QALY. Static downstream-case multipliers of 0.2 and 0.5 reduced the ICER to JPY 13.87 million and JPY 10.93 million per QALY, respectively, but did not cross the reference threshold. A 10% conditional hospitalization-risk scenario produced JPY 6.00 million per QALY.

The maximum total PEP intervention cost consistent with zero INMB was JPY 19,121 per contact. Holding other delivery costs fixed, the drug acquisition-cost threshold was JPY 11,021 per course, approximately 22% of the modeled current price. At current costs, the conditional hospitalization probability would need to reach approximately 10.6% for zero INMB. The post-acute QALY loss threshold was 0.1348 QALYs per surviving symptomatic case.

### 3.6 Verification and Input Traceability

All prespecified input checks and derived-input audits passed. Fifteen unit tests passed in a clean execution environment. Registered source values for hospitalization cost, mortality, length of stay, trial risks, and derived quantities were reproduced by dedicated traceability checks. Because several checks compare the model with its own selected input sources, they demonstrate correct transfer and internal consistency rather than independent predictive external validation. The full audit trail is provided in Online Resources 1-3.

## 4 Discussion

### 4.1 Principal Findings

Ensitrelvir post-exposure prophylaxis was projected to improve health but was unlikely to be cost-effective at current Japanese acquisition and delivery costs when evaluated at JPY 5 million per QALY. The ICER was approximately JPY 16.8 million per QALY. The probability of cost-effectiveness remained only 2.28% at JPY 10 million per QALY, and the conclusion remained unfavorable across conservative efficacy, post-acute utility, mortality, and hospitalization-attribution scenarios.

The result is driven by the economic structure of prophylaxis. Every eligible exposed contact incurs the intervention cost, whereas the trial-observed benefit is prevention of symptomatic disease. Hospitalization and death are uncommon and were absent in SCORPIO-PEP. Consequently, even a large relative reduction in symptomatic disease translates into limited expected savings from severe events. The principal modeled health benefit arose from avoided post-acute quality-of-life loss, which was also the largest structural uncertainty.

### 4.2 Interpretation for Policy and Practice

The findings do not imply that ensitrelvir PEP lacks clinical value. They indicate that routine prophylaxis for all high-risk household contacts is difficult to justify at the modeled price under a public payer perspective. The drug acquisition-cost threshold of approximately JPY 11,000 provides a transparent value-based pricing benchmark, although it is conditional on the model population, evidence sources, and JPY 5 million reference value.

Risk targeting may improve value. Exploratory scenarios for older or immunocompromised populations produced lower ICERs, but still exceeded the reference value. The largest improvement occurred in the B-cell-depleting therapy scenario, which remained approximately JPY 10.03 million per QALY. Even a 10% hospitalization probability conditional on symptomatic disease produced an ICER of approximately JPY 6.00 million per QALY. These scenarios use hospitalization multipliers from infected populations [14] rather than randomized PEP data and should be interpreted as hypothesis-generating.

Implementation costs also matter because prophylaxis must be initiated rapidly. Testing, clinical assessment, prescribing, dispensing, and delivery may be organized through face-to-face or telemedicine pathways. Lower-cost delivery alone did not reverse the conclusion, but delivery design becomes increasingly important if acquisition price is reduced. Static downstream-case bounds improved the ICER but did not make prophylaxis cost-effective. Because these calculations assigned downstream cases the same hospitalization and post-acute outcomes as the high-risk contact population, they should be interpreted as upper bounds rather than estimates from a dynamic transmission model.

### 4.3 Comparison with Other Economic Evidence

Previous Japanese economic evaluations of oral antivirals addressed treatment after diagnosis rather than prophylaxis and therefore applied drug costs to infected patients rather than exposed contacts [13]. This distinction materially changes the cost-effectiveness mechanism. Our breakthrough-treatment scenario jointly increased symptomatic-care cost and reduced hospitalization risk, and it left the conclusion essentially unchanged. Published Japanese studies also show continuing hospitalization burden among vulnerable populations and measurable post-acute quality-of-life loss [7, 8, 14, 15], supporting their inclusion in scenario and uncertainty analyses while underscoring the need for PEP-specific outcome data.

### 4.4 Strengths and Limitations

The model directly used randomized efficacy from a prespecified high-risk subgroup analysis and explicitly separated trial-observed symptomatic prevention from externally modeled severe outcomes. A conservative scenario replaced the subgroup relative risk with the overall-trial relative risk. The hospitalization source endpoint was identified as all-cause, and its uncertain attribution was represented as a structural parameter rather than treated as a hidden causal estimate.

Publication-critical inputs were derived from Japanese sources where possible, including hospitalization, inpatient mortality, hospital cost, EQ-5D-5L, population utility norms, life tables, drug price, and reimbursement schedules. Machine-readable parameter, evidence, extrapolation, and traceability registers allow each input to be followed from source to model. Price years and the absence of direct re-pricing for transferred cost studies are explicitly reported.

The analysis is fully reproducible in Python. It includes deterministic and probabilistic sensitivity analyses, efficacy, post-acute, breakthrough-treatment, risk, delivery, and static downstream-case scenarios, threshold analyses, independent recalculation of derived inputs, 15 unit tests, and a versioned public release archive. This analysis has several limitations. The pivotal trial observed no COVID-19-related hospitalization or death, so severe-outcome benefits are mediated model projections rather than randomized estimates. The hospitalization input was a 28-day all-cause admission endpoint among diagnosed high-risk outpatients, and the base case attributed all admissions to COVID-19. This may favor prophylaxis when admissions were incidental or unrelated. The conclusion nevertheless remained unfavorable when half or none of the admissions were attributed to COVID-19.

Hospitalization risk, in-hospital mortality, and hospital cost were linked across separate populations and calendar periods. The untreated outpatient cohort was younger and drawn largely from employment-based insurance, whereas the inpatient mortality cohort had a median age of 82 years and the cost study reflected a university hospital in 2021. This linkage may overstate cost and mortality per hospitalization and therefore favor prophylaxis. Removing mortality benefit or all hospitalization attribution did not change the decision. The post-acute utility input came from a retrospective online recall survey at one clinic, with 302 COVID-19 respondents, 77 test-negative controls, a young median age, no prospectively measured pre-infection baseline utility, and potential recall and response-selection bias [8]. Test-negative control utilities approached the ceiling in later intervals. Applying the full mean difference to all surviving symptomatic cases may therefore overestimate post-acute benefit. The decision remained unfavorable when the 6-12-month interval was removed, the integrated loss was halved, or post-acute benefit was excluded entirely.

The breakthrough-treatment and downstream-case analyses were exploratory. The treatment scenario assumed 50% uptake and observational treated and untreated hospitalization risks. Additional consultation, testing, and dispensing costs for treatment were not included; adding them would slightly favor prophylaxis by increasing avoided treatment costs but would not materially change the conclusion. The static model did not represent household contact networks, generation time, depletion of susceptible persons, variant-specific infectivity, or feedback from reduced infection. Downstream cases were assigned the same conditional outcomes as the high-risk contact population, so those scenarios are upper-bound calculations rather than dynamic transmission estimates.

Transferred outpatient and inpatient costs retained their source-year nominal values rather than being fully repriced to 2026. The FY2026 two-point outpatient inflation add-on, equivalent to JPY 20, was omitted because its effect is immaterial, and facility-dependent wage add-ons were excluded because eligibility varies. Future variants, waning immunity, antiviral resistance, productivity loss, caregiver effects, and broader societal costs were also omitted. Finally, no patients, caregivers, clinicians, or payers were directly involved in model design, which limits the model’s ability to reflect stakeholder preferences concerning access, testing burden, rapid delivery, and risk targeting.

## 5 Conclusions

Ensitrelvir post-exposure prophylaxis was projected to prevent symptomatic COVID-19 and generate QALY gains among high-risk household contacts in Japan, but it was unlikely to be cost-effective at current acquisition and delivery costs at JPY 5 million per QALY. The conclusion remained unfavorable after applying the overall-trial relative risk, reducing or removing post-acute utility benefit, removing modeled mortality, attributing no admissions to COVID-19, and allowing for post-diagnosis treatment. Value-based pricing and exceptionally high baseline risk are the main pathways toward improved economic value. Future studies should directly quantify severe outcomes, post-acute burden, transmission effects, and implementation costs in PEP-eligible populations.

## Supporting information

Model input and output data

## Data Availability

All data used in this study are published aggregate data and publicly available reimbursement information. All model inputs, analysis outputs, and the complete Python model are available online at https://github.com/toshtanig/ensitrelvir_PEP_CEA (release v0.3.2) and in the Supplementary Materials. No individual-level patient data were used.

## Acknowledgements

Not applicable.

## Declarations

### Ethics Approval

Ethics approval was not required because this model-based study used only published aggregate data and publicly available reimbursement information.

### Consent to Participate

Not applicable.

### Consent for Publication

Not applicable.

### Availability of Data and Material

All model inputs supporting the findings are supplied in Online Resource 2 and are publicly available with the versioned release at https://github.com/toshtanig/ensitrelvir_PEP_CEA. No individual-level patient data were used.

### Code Availability

The complete Python model version 0.3.2, tests, scenario definitions, environment files, and generated outputs are publicly available at https://github.com/toshtanig/ensitrelvir_PEP_CEA. The Git release tag v0.3.2 and the SHA-256 checksum of the release archive (2ad15256ae63f3ec445297b4ca0a6f4cd1ed35dc6c90bc45443187d2d2b33526) identify the computational record used for this analysis. The exact archive will be deposited in a permanent repository upon journal submission.

### Author Contributions

Toshibumi Taniguchi conceived the study, drafted the manuscript, and identified and obtained the model parameters. Misuzu Yahaba, Hiroshi Yoshikawa, and Hidetoshi Igari contributed to the clinical interpretation of the findings and critically revised the manuscript. Daisuke Sato provided health economic expertise and oversaw the accuracy and internal consistency of the cost-effectiveness model. Toshibumi Taniguchi is the guarantor. All authors read and approved the final manuscript.

## Supplementary Information

Online Resource 1: Supplementary methods, equations, evidence and extrapolation tables, sensitivity analyses, and additional figures (PDF).

Online Resource 2: Machine-readable model inputs, evidence registers, analysis outputs, and audit tables (XLSX).

Online Resource 3: Reproducible Python model, tests, environment specifications, source data, and generated outputs (ZIP).

