## Supplementary appendix for "Cost-Effectiveness of Ensitrelvir for COVID-19 Post-Exposure Prophylaxis Among High-Risk Household Contacts in Japan: A Model-Based Cost-Utility Analysis"

Journal: Applied Health Economics and Health Policy

Authors: Toshibumi Taniguchi, MD, PhD; Misuzu Yahaba, MD, PhD; Hiroshi Yoshikawa, MD; Hidetoshi Igari, MD, PhD; Daisuke Sato, PhD

Version 1.4, 15 September 2026; model version 0.3.2

### Contents


- S1 Decision problem and model equations
- S2 Evidence identification and extrapolation
- S3 Clinical effectiveness
- S4 Hospitalization, mortality, utilities, and costs
- S5 Sensitivity and threshold analyses
- S6 Verification, input traceability, and reproducibility
- Supplementary Tables S1-S11
- Supplementary Figures S1-S2


### S1 Decision Problem and Model Equations

The model compares a five-day ensitrelvir post-exposure prophylaxis course with no pharmacologic prophylaxis among high-risk household contacts in Japan. The core causal assumption is that ensitrelvir changes the probability of symptomatic COVID-19 but does not directly change the conditional severity of breakthrough symptomatic infection.

For strategy j, expected symptomatic disease is P(Symptomatic | j). COVID-19-attributable hospitalization is P(Symptomatic | j) x P(Hospitalization | Symptomatic) x AF, where AF is the hospitalization-attribution fraction. Death is the attributable hospitalization probability multiplied by P(Death | Hospitalization).

Expected cost equals intervention cost plus symptomatic outpatient cost, hospitalization cost, post-acute direct medical cost, and incremental adverse-event cost. Expected QALY loss equals acute morbidity loss, acute mortality loss, post-acute morbidity loss, and incremental adverse-event loss. Incremental QALYs are no-prophylaxis QALY loss minus ensitrelvir QALY loss. INMB equals willingness-to-pay multiplied by incremental QALYs minus incremental cost.

The model uses a 1-year horizon for post-acute morbidity and a lifetime horizon for acute mortality. Drug acquisition and PEP-delivery inputs use 2026 prices and fee schedules. Transferred outpatient and inpatient costs retain their source-year nominal values and are varied using cost multipliers. Costs are incurred within 1 year and are not materially affected by discounting. Future QALYs lost through acute death are discounted at 2% annually.

**Table S1 Decision problem defined using PICOTS**

| Element | Specification |
|---|---|
| Population | Household contacts with at least one risk factor for severe COVID-19, eligible within 72 hours of index-case symptom onset |
| Intervention | Ensitrelvir 375 mg on day 1 and 125 mg on days 2-5 |
| Comparator | No pharmacologic post-exposure prophylaxis |
| Perspective | Japanese public healthcare payer |
| Outcomes | Costs, QALYs, ICER, INMB, symptomatic disease, hospitalization, death |
| Time horizon | Acute episode plus 1-year post-acute morbidity and lifetime mortality QALY loss |
| Setting | Japan; household exposure |
| Price year | 2026 JPY for drug and PEP delivery; 2023 and 2021 nominal source-year values for transferred outpatient and inpatient costs, tested with 0.8-1.2 multipliers |

Abbreviations: COVID-19 coronavirus disease 2019; PEP post-exposure prophylaxis; PICOTS population, intervention, comparator, outcomes, timing, and setting; QALY quality-adjusted life-year.

### S2 Evidence Identification and Extrapolation

Targeted searches were performed for ensitrelvir post-exposure prophylaxis efficacy; Japanese hospitalization and mortality; acute and post-acute EQ-5D-5L; hospital, outpatient, and post-acute cost; life tables; population utility; and current reimbursement. Evidence was prioritized in the following order: randomized trial for efficacy, Japanese contemporary observational data for baseline outcomes, Japanese patient-level cost and utility studies, official price and fee schedules, and explicit structural assumptions when empirical data were unavailable.

Each source-to-target transformation was recorded in a separate extrapolation register. Important differences included the use of all-cause rather than COVID-19-specific hospitalization, separate populations for outpatient risk and inpatient mortality, a 2021 university-hospital cost cohort, and a younger post-acute utility survey than the high-risk target population. Comparisons against source values are described as input traceability or face-validity checks rather than independent predictive external validation.

**Table S2 Selected evidence sources and limitations**

| ID | Design/source | Model use | Principal limitation |
|---|---|---|---|
| E01 | Phase 3 double-blind randomized placebo-controlled trial | Arm-specific symptomatic COVID-19 risk; overall all-infection risk; safety; high-risk subgroup efficacy. | No COVID-19-related hospitalizations or deaths; downstream severe outcomes cannot be estimated directly. Highly immune trial population. |
| E02 | Official regulatory and price announcement | Approved prevention indication, dose, tablet price, course acquisition cost. | Manufacturer source; drug price should be rechecked against the official NHI price list at analysis lock. |
| E03 | Official methodological guideline | Perspective, QALY outcome, Japanese utility preference, costing, discounting, modeling, reproducibility, scenario and probabilistic sensitivity analysis. | English version is a translation; Japanese version prevails if discrepancies arise. |
| E08 | Single-center claims-based hospital cost study | Hospitalization cost input and source-traceability comparator. | Single-center 2021 university-hospital cohort, including pre-Omicron practice; nominal source-year cost was not fully re-priced to 2026. |
| E12 | Multicenter retrospective hospital database study | Contemporary hospitalized-case age, sex, length of stay, ICU use, ventilation, and in-hospital mortality; late-Omicron median age 82 and overall male share 54% inform the death-QALY proxy. | Hospitalized population is selected and older than the PEP trial. Age and sex of admissions, rather than decedents, are used as a proxy; the study was industry sponsored. |
| E13 | Official national life table | Age-sex survival probabilities for lifetime QALY loss after acute death. | Life-table input is truncated at age 105 in the repository; residual survival beyond 105 is omitted. |
| E14 | Retrospective online recall survey with a test-negative comparison group | Acute utility decrement and direct integration of post-acute utility differences from months 1 to 12. | Single Tokyo clinic; 302 COVID-19 respondents and 77 controls; median age 42; 28.3% response; retrospectively recalled intervals; no prospectively measured baseline; recall and response-selection bias; near-ceiling later control utilities. |

| ID | Design/source | Model use | Principal limitation |
|---|---|---|---|
| E17 | Retrospective employer-based claims cohort | Untreated high-risk outpatient 28-day all-cause hospitalization risk and transportability scenarios. | Employer-based claims population with few adults aged 65 years or older; vaccination status unavailable; primary endpoint was all-cause hospitalization; exposure was antiviral treatment after diagnosis, not PEP. |
| E18 | Official product label | Prevention dose and requirement to start within 72 hours after contact; implementation scenario definition. | The trial required baseline negativity, while the label does not state that a negative test is mandatory. Local practice may differ. |
| E21 | Retrospective claims cohort | Exploratory immunocompromised, B-cell-depleting therapy, and hematologic-malignancy hospitalization-rate multipliers. | Population incidence-rate ratios are transferred to hospitalization conditional on symptomatic infection. Follow-up was calendar year 2023 and several authors were employed by the sponsor. |

Abbreviations: COVID-19 coronavirus disease 2019; EQ-5D-5L EuroQol 5-Dimension 5-Level; NHI National Health Insurance; PEP post-exposure prophylaxis; QALY quality-adjusted life-year.

### S3 Clinical Effectiveness

The high-risk subgroup supplied arm-specific symptomatic risks. The absolute risk reduction was 0.09893 - 0.02356 = 0.07537. The number needed to treat was 1/0.07537 = 13.27. The trial reported that subgroup analyses were prespecified. A conservative scenario applied the overall-trial relative risk of 0.33 to the high-risk untreated risk. In PSA, untreated risk was sampled from a Jeffreys beta posterior and combined with a log-normal relative risk. This parameterization preserves uncertainty in baseline risk and relative treatment effect but assumes zero covariance because the joint estimate was unavailable.

**Table S3 Trial event counts used in the high-risk base case**

| Arm | Symptomatic events | Participants | Risk |
|---|---|---|---|
| Ensitrelvir | 9 | 382 | 0.02356 |
| No prophylaxis | 37 | 374 | 0.09893 |

Abbreviation: None.

### S4 Hospitalization, Mortality, Utilities, and Costs

Hospitalization probability was based on untreated high-risk outpatients in a retrospective claims study whose endpoint was 28-day all-cause hospitalization. A separate attribution factor was necessary because the endpoint was not COVID-19-specific. The base case uses 1.0 to avoid arbitrarily removing potentially related admissions, while 0.5 and 0.0 quantify structural optimism. In-hospital mortality and inpatient cost came from separate populations and calendar periods. A breakthrough-treatment scenario jointly added antiviral acquisition cost for 50% of symptomatic cases and used the corresponding treated and untreated hospitalization risks reported in the claims study. It did not add separate consultation, testing, or dispensing costs for post-diagnosis treatment; including those costs would slightly favor prophylaxis.

Remaining QALYs lost per death were calculated by integrating annual survival and age-sex population utility, weighted by the proxy inpatient cohort distribution. The post-acute QALY loss was calculated from a retrospective online recall survey at one Tokyo clinic with 302 COVID-19 respondents and 77 test-negative controls. Participants retrospectively reported EQ-5D-5L for the acute period and months 1-3, 3-6, and 6-12. The base-case integral was 0.03225 QALYs per surviving symptomatic case. Conservative scenarios removed the 6-12-month interval, halved the total loss, or removed post-acute utility benefit. General-population norms were not substituted for symptomatic test-negative controls because this would not preserve like-for-like symptom and selection characteristics. The 7-day acute decrement is followed by no additional acute decrement during days 8-30, which is conservative for prophylaxis. Both principal derivations are recalculated by audit scripts.

**Table S4 FY2026 post-exposure prophylaxis delivery microcost**

| ID | Component | Points | JPY | Included | Notes |
|---|---|---|---|---|---|
| M01 | Initial consultation | 291 | 2,910 | yes | Face-to-face initial visit. Telemedicine is a scenario. The FY2026 two-point outpatient inflation add-on (JPY 20) was excluded because its effect is immaterial. Facility-dependent wage add-ons were separately excluded because eligibility varies. |
| M02 | SARS-CoV-2 qualitative antigen test | 150 | 1,500 | yes | Included as an implementation pathway assumption, not a labeled mandatory test. |
| M03 | Immunological test interpretation fee | 144 | 1,440 | yes | Assumes no other immunological test interpretation fee billed in the month. |
| M04 | Nasal or pharyngeal swab collection | 25 | 250 | yes | Specimen collection fee. |
| M05 | Prescription fee | 60 | 600 | yes | Standard prescription fee for a non-complex prescription. |
| P01 | Pharmacy basic dispensing fee 1 | 47 | 470 | yes | Representative community-pharmacy base fee; actual pharmacy category can differ. |
| P02 | Oral drug preparation fee | 24 | 240 | yes | Five-day oral course. |
| P03 | Dispensing management fee, less than 28 days | 10 | 100 | yes | Short-course oral medication. |
| P04 | Medication management and counselling, new or other patient | 59 | 590 | yes | One-off PEP dispensing; repeat-with-notebook fee is lower. |

| ID | Component | Points | JPY | Included | Notes |
|---|---|---|---|---|---|
| NOTE | FY2026 add-ons not included in the base microcost | | | no | The two-point outpatient inflation add-on is JPY 20 and immaterial. Facility-dependent wage add-ons were excluded because eligibility varies; uncertainty is covered by delivery-cost ranges. |

Abbreviations: FY fiscal year; JPY Japanese yen; SARS-CoV-2 severe acute respiratory syndrome coronavirus 2.

Table S5 Post-acute QALY integration from a Japanese retrospective recall EQ-5D-5L survey

| Interval | Start | End | COVID utility | Control utility | Difference | QALY loss |
|---|---|---|---|---|---|---|
| PA01 | 1 | 3 | 0.907 | 0.976 | 0.069 | 0.0115 |
| PA02 | 3 | 6 | 0.961 | 0.994 | 0.033 | 0.00825 |
| PA03 | 6 | 12 | 0.97 | 0.995 | 0.025 | 0.0125 |

Abbreviations: COVID-19 coronavirus disease 2019; QALY quality-adjusted life-year.

**Table S6 Proxy cohort used to calibrate remaining QALYs lost per acute death**

| Age | Sex | Weight | Mortality HR | Source | Notes |
|---|---|---|---|---|---|
| 82 | male | 0.54 | 1.0 | E12 | Late-Omicron Japanese hospitalized cohort median age 82; male share 54% from the overall contemporary inpatient cohort is used as a proxy for acute COVID-19 deaths. |
| 82 | female | 0.46 | 1.0 | E12 | Female share inferred as 1 minus the reported 54% male share; age and sex are proxies because decedent-specific distribution was unavailable. |

Abbreviation: HR hazard ratio.

**Table S7 Complete model parameter register**

| Parameter ID | Description | Base | Low | High | Unit | PSA distribution | Source | Grade |
|---|---|---|---|---|---|---|---|---|
| discount_rate_cost | Annual discount rate for costs | 0.02 | 0 | 0.04 | proportion/year | pert | E03 | A |
| discount_rate_qaly | Annual discount rate for QALYs | 0.02 | 0 | 0.04 | proportion/year | pert | E03 | A |

| Parameter ID | Description | Base | Low | High | Unit | PSA distribution | Source | Grade |
|---|---|---|---|---|---|---|---|---|
| wtp_per_qaly | Willingness-to-pay per QALY | 5,000,000 | 5,000,000 | 5,000,000 | JPY/QALY | fixed | E03 | B |
| pcc_horizon_months | PCC module horizon | 120 | 60 | 600 | months | fixed | A01 | D |
| drug_cost_pep | Ensitrelvir acquisition cost per PEP course | 49,630 | 0 | 49,630 | JPY/course | fixed | E02 | A |
| administration_cost_pep | Oral antiviral administration cost | 8,100 | 4,530 | 10,000 | JPY/course | pert | E20 | A |
| additional_pep_access_cost | Additional PEP access and dispensing cost | 0 | 0 | 10,000 | JPY/course | fixed | A02 | D |
| p_hospital_given_symptomatic | Hospitalization probability conditional on symptomatic COVID-19 | 0.00785 | 0.00494 | 0.01298 | proportion | pert | E17 | B |
| hospitalization_attribution_fraction | Fraction of observed all-cause hospitalizations attributable to COVID-19 | 1 | 0 | 1 | proportion | fixed | A11 | D |
| p_death_given_hospital | In-hospital mortality conditional on COVID-19 hospitalization | 0.12 | 0.06 | 0.2 | proportion | pert | E12 | B |
| p_death_outpatient | Mortality probability among nonhospitalized symptomatic cases | 0 | 0 | 0 | proportion | fixed | A10 | C |
| p_icu_given_hospital | ICU admission probability conditional on hospitalization | 0.107 | 0.05 | 0.2 | proportion | pert | E04 | C |

| Parameter ID | Description | Base | Low | High | Unit | PSA distribution | Source | Grade |
|---|---|---|---|---|---|---|---|---|
| p_mv_given_icu | Mechanical ventilation probability conditional on ICU admission | 0.6454 | 0.4 | 0.8 | proportion | pert | E04 | C |
| p_death_ward | Inpatient mortality in general ward | 0.0319 | 0.01 | 0.08 | proportion | pert | E04 | C |
| p_death_icu_no_mv | Mortality in ICU without mechanical ventilation | 0.118 | 0.05 | 0.25 | proportion | pert | E04 | C |
| p_death_icu_mv | Mortality in ICU with mechanical ventilation | 0.118 | 0.05 | 0.3 | proportion | pert | E04 | C |
| symptom_duration_outpatient_days | Duration of acute symptoms in nonhospitalized cases | 7 | 4 | 14 | days | lognormal_cv | E04 | C |
| disutility_outpatient | Utility decrement for nonhospitalized symptomatic COVID-19 | 0.258 | 0.181 | 0.335 | utility decrement | pert | E14 | B |
| disutility_hospital | Utility decrement during COVID-19 hospitalization | 0.284 | 0.056 | 0.5 | utility decrement | pert | E07 | C |
| disutility_ward | Utility decrement for general ward COVID-19 | 0.056 | 0.02 | 0.15 | utility decrement | pert | E07 | B |
| disutility_icu | Utility decrement for ICU COVID-19 | 0.284 | 0.15 | 0.5 | utility decrement | pert | E07 | B |
| remaining_qaly_if_acute_death | Discounted remaining QALYs lost per acute COVID-19 death | 6.46949 | 5.320961 | 9.778527 | QALY/ death | pert | E19 | B |
| cost_outpatient | Outpatient COVID-19 treatment cost | 26,927 | 21,542 | 32,312 | JPY/case | gamma_cv | E04 | B |

| Parameter ID | Description | Base | Low | High | Unit | PSA distribution | Source | Grade |
|---|---|---|---|---|---|---|---|---|
| cost_hospitalization | Direct medical cost per COVID-19 hospitalization | 1,304,431 | 798,810 | 6,210,607 | JPY/ admission | pert | E08 | B |
| los_hospital_days | Length of stay for COVID-19 hospitalization | 21 | 10 | 31 | days | pert | E12 | B |
| acute_cost_price_adjustment_factor | Acute cost price and practice adjustment factor | 1 | 0.8 | 1.2 | multiplier | pert | A08 | D |
| cost_ward_per_day | General ward cost per day | 71,515 | 57,212 | 85,818 | JPY/day | gamma_cv | E04 | B |
| cost_icu_no_mv_per_day | ICU without mechanical ventilation cost per day | 193,598 | 154,878 | 232,318 | JPY/day | gamma_cv | E04 | B |
| cost_icu_mv_per_day | ICU with mechanical ventilation cost per day | 324,254 | 259,403 | 389,105 | JPY/day | gamma_cv | E04 | B |
| los_ward_days | Length of stay in general ward | 8.76 | 7 | 10.5 | days | lognormal_cv | E04 | B |
| los_icu_no_mv_days | Length of stay in ICU without mechanical ventilation | 10.9 | 8 | 14 | days | lognormal_cv | E04 | B |
| los_icu_mv_days | Length of stay in ICU with mechanical ventilation | 9.78 | 7 | 14 | days | lognormal_cv | E04 | B |
| postacute_qaly_loss_per_surviving_symptomatic_case | One-year post-acute QALY loss per surviving symptomatic COVID-19 case | 0.03225 | 0.010667 | 0.052 | QALY/ case | pert | E14 | B |
| postacute_cost_per_surviving_symptomatic_case | Post-acute direct medical cost per surviving symptomatic COVID-19 case | 0 | 0 | 200,000 | JPY/case | fixed | A07 | D |

| Parameter ID | Description | Base | Low | High | Unit | PSA distribution | Source | Grade |
|---|---|---|---|---|---|---|---|---|
| p_pcc_outpatient | PCC probability after nonhospitalized symptomatic COVID-19 | 0.063 | 0.02 | 0.118 | proportion | pert | E05 | B |
| pcc_hospital_multiplier | PCC risk multiplier after hospitalization | 2 | 1 | 5 | risk multiplier | pert | A04 | D |
| p_pcc_asymptomatic | PCC probability after asymptomatic infection | 0 | 0 | 0.03 | proportion | fixed | A05 | D |
| pcc_mean_duration_months | Mean duration of PCC | 12 | 6 | 60 | months | lognormal_cv | A06 | D |
| disutility_pcc | Utility decrement for PCC | 0.027 | 0.01 | 0.1 | utility decrement | pert | E06 | B |
| cost_pcc_annual | Annual direct medical cost of PCC | 0 | 0 | 200,000 | JPY/person-year | fixed | A07 | D |
| incremental_ae_cost_pep | Incremental adverse-event cost attributable to PEP | 0 | 0 | 5,000 | JPY/course | fixed | E01 | C |
| incremental_ae_qaly_loss_pep | Incremental adverse-event QALY loss attributable to PEP | 0 | 0 | 0.0005 | QALY/course | fixed | E01 | C |
| secondary_symptomatic_case_multiplier | Additional downstream symptomatic cases per direct symptomatic case | 0 | 0 | 0.5 | cases/case | fixed | A12 | D |

Abbreviations: JPY Japanese yen; PCC post-COVID condition; PSA probabilistic sensitivity analysis; QALY quality-adjusted life-year.

### S5 Sensitivity, Scenario, and Threshold Analyses

One-way sensitivity analysis varied one parameter at a time over the registered range and recalculated INMB. The main manuscript tornado plot excludes drug acquisition cost because its full 0-100% variation would dominate the display and price is evaluated separately by threshold analysis; the complete DSA table below retains drug cost. Structural analyses were not assigned arbitrary probabilistic distributions when no empirical basis existed. PERT-type distributions were used when lower, modal, and upper bounds were available without an empirical parametric fit.

Probabilistic sensitivity analysis used 10,000 draws and seed 20260825. The complete draws are included in Online Resource 3. Threshold analyses used parameter grids and interpolation to identify the zero-INMB crossing when one existed within the prespecified range. Additional conservative scenarios applied the overall-trial relative risk, shortened or reduced post-acute utility loss, incorporated 50% breakthrough-treatment uptake, and added static downstream symptomatic-case multipliers of 0.2 and 0.5. Downstream cases were assigned the same conditional hospitalization and post-acute outcomes as the high-risk contact population, so these are upper-bound calculations rather than dynamic transmission models. The 10% conditional hospitalization scenario is hypothetical and was not observed in any Japanese source used in the model.

**Table S8 Complete scenario analysis**

| Scenario ID | Description | Incremental cost | Incremental QALY | ICER, JPY m/QALY | CE at 5m |
|---|---|---|---|---|---|
| base_high_risk | Publication base case for high-risk household contacts using Japanese hospitalization, inpatient mortality, life-table QALYs, and a directly integrated one-year post-acute EQ-5D profile. | 54,944.67 | 0.003267 | 16.82 | False |
| high_risk_no_postacute | Structural sensitivity analysis excluding all post-acute QALY loss and direct medical cost. | 54,944.67 | 0.000839 | 65.51 | False |
| high_risk_no_mortality | Structural sensitivity analysis excluding acute mortality benefit. | 54,944.67 | 0.002810 | 19.55 | False |
| high_risk_symptom_only | Lower-bound scenario limited to outpatient symptomatic illness and outpatient cost. | 55,700.51 | 0.000373 | 149.46 | False |
| high_risk_face_to_face_no_test | Face-to-face consultation and dispensing without separately billed pre-PEP testing. | 51,754.67 | 0.003267 | 15.84 | False |
| high_risk_telemedicine_no_test | Telemedicine consultation and dispensing without separately billed pre-PEP testing. | 51,374.67 | 0.003267 | 15.72 | False |
| high_risk_telemedicine_with_test | Telemedicine consultation plus antigen testing and dispensing. | 54,564.67 | 0.003267 | 16.70 | False |
| high_risk_age65_hospital_risk | Uses the 1.298% hospitalization risk observed among untreated patients aged 65 years or older in the JMDC study. | 54,450.73 | 0.003570 | 15.25 | False |

| Scenario ID | Description | Incremental cost | Incremental QALY | ICER, JPY m/QALY | CE at 5m |
|---|---|---|---|---|---|
| high_risk_old_model_hospital_risk | Uses the 1.77% hospitalization risk from the earlier Japanese treatment CEA. | 53,996.26 | 0.003849 | 14.03 | False |
| high_risk_immunocompromised_rr26 | Exploratory immunocompromised scenario applying a 2.60 hospitalization-rate multiplier to the base risk. | 53,735.33 | 0.004009 | 13.40 | False |
| high_risk_postacute_cost_50k | Adds JPY 50,000 post-acute direct medical cost per surviving symptomatic case. | 51,179.72 | 0.003267 | 15.67 | False |
| high_risk_postacute_cost_100k | Adds JPY 100,000 post-acute direct medical cost per surviving symptomatic case. | 47,414.77 | 0.003267 | 14.51 | False |
| high_risk_postacute_cost_200k | Adds JPY 200,000 post-acute direct medical cost per surviving symptomatic case. | 39,884.87 | 0.003267 | 12.21 | False |
| legacy_incidence_duration | Structural sensitivity analysis retaining the v0.1 ICU, ventilation, and PCC incidence-duration model. | 54,591.15 | 0.001614 | 33.83 | False |
| very_high_risk | Hypothetical 10% conditional hospitalization-risk scenario; the value was not observed in any Japanese source used in the model. | 46,071.96 | 0.007672 | 6.00 | False |
| overall_exploratory | Exploratory overall-trial population analysis with a lower hospitalization risk. | 55,701.81 | 0.002815 | 19.79 | False |
| overall_any_infection_pcc | Legacy exploratory model using the all-infection endpoint and PCC after asymptomatic infection. | 55,837.55 | 0.000874 | 63.89 | False |

| Scenario ID | Description | Incremental cost | Incremental QALY | ICER, JPY m/QALY | CE at 5m |
|---|---|---|---|---|---|
| high_risk_hospital_attribution_50pct | Structural sensitivity analysis attributing 50% of observed all-cause hospitalizations to COVID-19. | 55,322.59 | 0.003035 | 18.23 | False |
| high_risk_hospital_attribution_0pct | Structural sensitivity analysis attributing none of the all-cause hospitalizations to COVID-19. | 55,700.51 | 0.002803 | 19.87 | False |
| high_risk_malignancy_or3941 | Exploratory malignancy scenario using the Japanese outpatient odds ratio of 3.941. | 52,788.96 | 0.004590 | 11.50 | False |
| high_risk_renal_or2003 | Exploratory renal-disease scenario using the Japanese outpatient odds ratio of 2.003. | 54,198.39 | 0.003725 | 14.55 | False |
| high_risk_immunosuppressive_or1637 | Exploratory immunosuppressive-drug scenario using the Japanese outpatient odds ratio of 1.637. | 54,469.36 | 0.003559 | 15.31 | False |
| high_risk_bcell_depleting_airr510 | Exploratory B-cell-depleting-therapy scenario applying a hospitalization aIRR of 5.10. | 51,845.72 | 0.005169 | 10.03 | False |
| high_risk_death_qaly_age75 | Mortality-QALY scenario calibrated to an age-75 proxy cohort. | 54,944.67 | 0.003502 | 15.69 | False |
| high_risk_death_qaly_age85 | Mortality-QALY scenario calibrated to an age-85 proxy cohort. | 54,944.67 | 0.003186 | 17.25 | False |
| high_risk_overall_rr033 | Conservative efficacy scenario applying the overall-trial risk ratio of 0.33 to the high-risk untreated risk. | 55,280.48 | 0.002873 | 19.24 | False |
| high_risk_postacute_through6m | Conservative post-acute scenario excluding the recalled utility difference from months 6 to 12. | 54,944.67 | 0.002326 | 23.62 | False |

| Scenario ID | Description | Incremental cost | Incremental QALY | ICER, JPY m/QALY | CE at 5m |
|---|---|---|---|---|---|
| high_risk_postacute_half | Conservative attenuation scenario using 50% of the base-case post-acute QALY loss. | 54,944.67 | 0.002053 | 26.76 | False |
| high_risk_breakthrough_treatment_50pct | Exploratory internally consistent scenario with 50% ensitrelvir treatment uptake after symptomatic breakthrough, adding acquisition cost and using a weighted hospitalization risk. | 53,226.42 | 0.003181 | 16.73 | False |
| high_risk_secondary_cases_0p2 | Exploratory static bounding analysis with 0.2 additional downstream symptomatic cases per direct symptomatic case. | 54,387.61 | 0.003921 | 13.87 | False |
| high_risk_secondary_cases_0p5 | Exploratory static bounding analysis with 0.5 additional downstream symptomatic cases per direct symptomatic case. | 53,552.01 | 0.004901 | 10.93 | False |

Abbreviations: CE cost-effective; EQ-5D EuroQol 5-Dimension; ICER incremental cost-effectiveness ratio; JPY Japanese yen; QALY quality-adjusted life-year.

### Table S9 One-way sensitivity analysis ranked by INMB span

| Parameter | Base | Low | High | Minimum INMB | Maximum INMB | Span |
|---|---|---|---|---|---|---|
| drug_cost_pep | 49,630 | 0 | 49,630 | -38,609.05 | 11,020.95 | 49,630 |
| postacute_qaly_loss_per_surviving_symptomatic_case | 0.03225 | 0.010667 | 0.052 | -46,735.07 | -31,173.27 | 15,561.79 |
| postacute_cost_per_surviving_symptomatic_case | 0 | 0 | 200,000 | -38,609.05 | -23,549.25 | 15,059.8 |
| trial_control_symptomatic_risk | 0.09893 | 0.071762 | 0.132274 | -43,893.73 | -32,226.5 | 11,667.23 |
| additional_pep_access_cost | 0 | 0 | 10,000 | -48,609.05 | -38,609.05 | 10,000 |
| trial_relative_risk_symptomatic_covid | 0.24 | 0.12 | 0.49 | -44,930.03 | -35,643.77 | 9,286.26 |
| administration_cost_pep | 8,100 | 4,530 | 10,000 | -40,509.05 | -35,039.05 | 5,470 |
| incremental_ae_cost_pep | 0 | 0 | 5,000 | -43,609.05 | -38,609.05 | 5,000 |

| Parameter | Base | Low | High | Minimum INMB | Maximum INMB | Span |
|---|---|---|---|---|---|---|
| cost_hospitalization | 1,304,431 | 798,810 | 6,210,607 | -38,908.2 | -35,706.29 | 3,201.91 |
| p_hospital_given_symptomatic | 0.00785 | 0.00494 | 0.01298 | -39,748.84 | -36,599.73 | 3,149.11 |
| hospitalization_attribution_fraction | 1 | 0 | 1 | -41,683.74 | -38,609.05 | 3,074.69 |
| p_death_given_hospital | 0.12 | 0.06 | 0.2 | -39,751.64 | -37,085.6 | 2,666.04 |
| symptom_duration_outpatient_days | 7 | 4 | 14 | -39,401.36 | -36,760.32 | 2,641.04 |
| incremental_ae_qaly_loss_pep | 0 | 0 | 0.0005 | -41,109.05 | -38,609.05 | 2,500 |
| remaining_qaly_if_acute_death | 6.46949 | 5.320961 | 9.778527 | -39,016.77 | -37,434.37 | 1,582.4 |
| acute_cost_price_adjustment_factor | 1 | 0.8 | 1.2 | -39,166.12 | -38,051.99 | 1,114.13 |
| disutility_outpatient | 0.258 | 0.181 | 0.335 | -39,160.8 | -38,057.3 | 1,103.51 |
| cost_outpatient | 26,927 | 21,542 | 32,312 | -39,011.73 | -38,206.37 | 805.362781 |
| disutility_hospital | 0.284 | 0.056 | 0.5 | -38,647.83 | -38,572.31 | 75.517954 |
| los_hospital_days | 21 | 10 | 31 | -38,634.35 | -38,586.05 | 48.304277 |
| discount_rate_cost | 0.02 | 0 | 0.04 | -38,609.05 | -38,609.05 | 0 |
| discount_rate_qaly | 0.02 | 0 | 0.04 | -38,609.05 | -38,609.05 | 0 |

Abbreviation: INMB incremental net monetary benefit.

**Table S10 Threshold analyses**

| Parameter | Zero-INMB crossing | Grid minimum | Grid maximum |
|---|---|---|---|
| drug_cost_pep | 11,020.95 | 0 | 60,000 |
| administration_cost_pep | No crossing | 0 | 15,000 |
| p_hospital_given_symptomatic | 0.106423 | 0 | 0.2 |
| hospitalization_attribution_fraction | No crossing | 0 | 1 |
| p_death_given_hospital | No crossing | 0 | 0.5 |
| remaining_qaly_if_acute_death | No crossing | 0 | 25 |
| postacute_qaly_loss_per_surviving_symptomatic_case | 0.134799 | 0 | 0.15 |
| postacute_cost_per_surviving_symptomatic_case | No crossing | 0 | 500,000 |

Abbreviation: INMB incremental net monetary benefit.

### S6 Verification, Input Traceability, and Reproducibility

Verification comprised input-range checks, trial arithmetic, source foreign-key checks, scenario execution, and unit tests. Derived inputs were independently recomputed from source tables. Registered source values for cost, length of stay, mortality, trial risks, and derived quantities were used for input traceability and face-validity checks. Because several checks compare the model with the same sources used to define inputs, they are not independent predictive external validation.

Table S11 Input traceability and face-validity checks

| ID | Metric | Modeled | Target | Unit | Interpretation |
|---|---|---|---|---|---|
| V01 | Median total inpatient cost, all hospitalized cases | 1,304,431 | 1,304,431 | JPY/admission | Model expectation is a mean-like pathway-weighted value; the external target is a median. |
| V02 | Median severe inpatient cost | 3,171,204.12 | 6,210,607 | JPY/admission | The external target is a median for severe 2021 cases and may include a longer or more resource-intensive pathway. |
| V03 | Median length of stay, all hospitalized cases | 21 | 10 | days | Historical single-center LOS comparator; the publication base uses the contemporary Japanese cohort instead. |
| V04 | Median length of stay, contemporary hospitalized cohort | 21 | 21 | days | Direct check against the contemporary Japanese cohort median LOS. |
| V05 | In-hospital mortality, contemporary hospitalized cohort | 0.12 | 0.12 | proportion | Direct check against contemporary Japanese in-hospital mortality. |

Abbreviations: JPY Japanese yen; LOS length of stay.

All input checks and three derived-input audits passed. Fifteen Python tests passed. The complete environment specification is included in Online Resource 3. The exact public release of model version 0.3.2 is available at https://github.com/toshtanig/ensitrelvir_PEP_CEA and archived at Zenodo under https://doi.org/10.5281/zenodo.22107356. The Git release tag and Zenodo DOI identify the computational record; a separate checksum citation is not required.

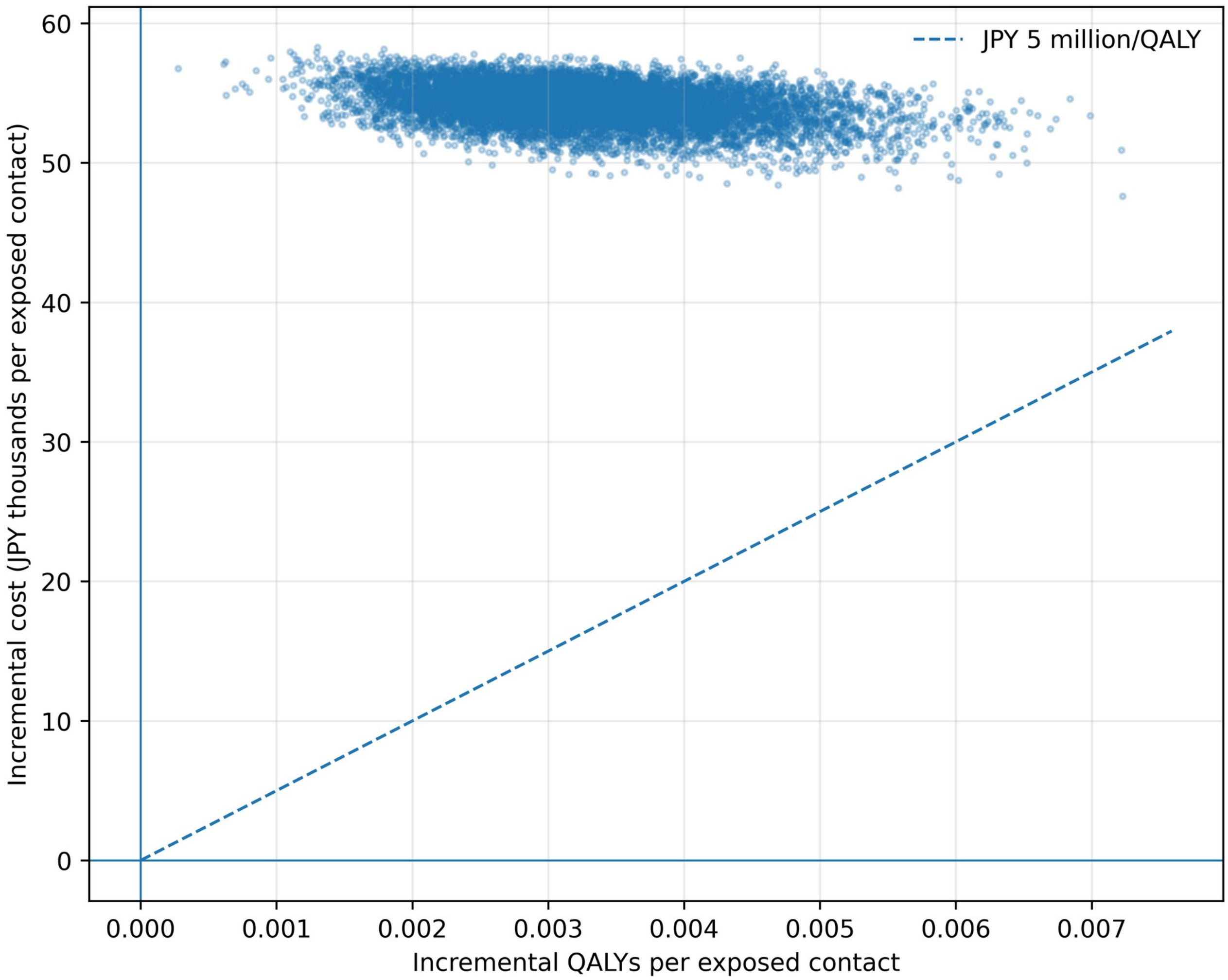


**Fig. S1** Probabilistic cost-effectiveness plane. The dashed line represents JPY 5 million per QALY. Abbreviations: JPY Japanese yen; QALY quality-adjusted life-year

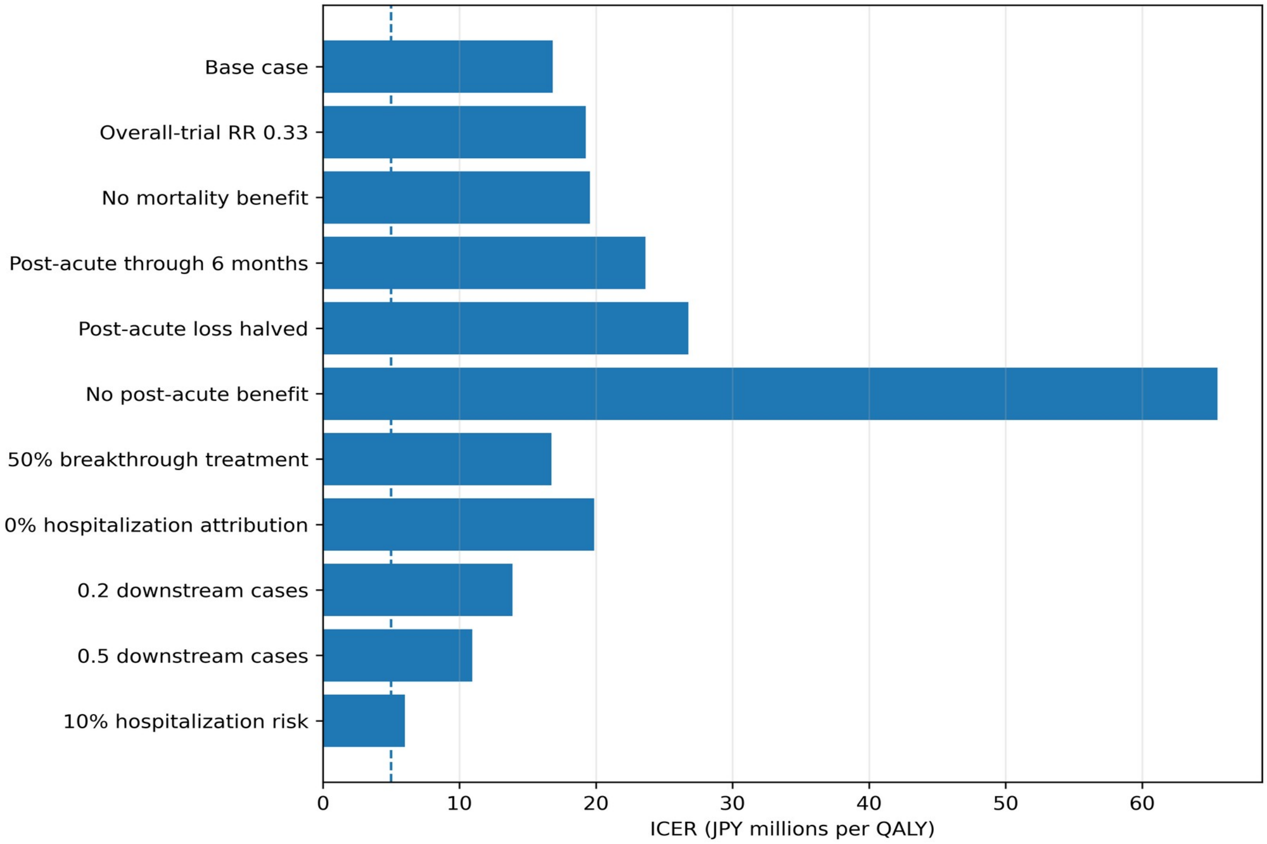


**Fig. S2** Selected conservative, structural, and exploratory scenario ICERs. The dashed vertical line represents JPY 5 million per QALY. Abbreviations: ICER incremental cost-effectiveness ratio; JPY Japanese yen; QALY quality-adjusted life-year
